# Anthracycline-mediated cardiac dysfunction: An endothelial perspective

**DOI:** 10.64898/2026.03.02.26347478

**Authors:** Kasturi Markandran, Tricia Jingwen Ng, Elena Tan, Czaryna Kaye Mendoza Clemente, Rina Miao Qin Wang, Yee Phong Lim, Karan Attal, Kristine Nicole Mendoza Clemente, Hannah Su-Ann Wee, S Hammresh, Christine Cheung, Sik Yin Roger Foo, Ching Kit Chen

**Affiliations:** Department of Paediatrics, Yong Loo Lin School of Medicine, National University of Singapore, Singapore; Cardiovascular Disease Translational Research Programme, Yong Loo Lin School of Medicine, National University of Singapore, Singapore; Cardiovascular Research Institute, Department of Medicine, Yong Loo Lin School of Medicine, National University of Singapore, National University Health System, Singapore, Singapore; Royal College of Surgeons in Ireland (Medicine), Dublin, Ireland; Lee Kong Chian School of Medicine, Experimental Medicine Building, 59 Nanyang Drive, Nanyang Technological University, Singapore 636921, Singapore; Institute of Molecular and Cell Biology, 61 Biopolis Drive, Proteos, Singapore 138673, Singapore; Division of Cardiology, Department of Paediatrics, Khoo Teck Puat–National University Children’s Medical Institute, National University Health System, Singapore

**Keywords:** Circulating endothelial cells, Endothelial progenitor cells, Endothelial injury, Senescence, Leukemia, Lymphoma

## Abstract

**Background:** Anthracyclines are central to childhood cancer therapy but predispose patients to cardiotoxicity leading to long-term cardiovascular risk. Endothelial injury and impaired repair contribute to this, yet pediatric data remain limited.

**Objective:** To longitudinally assess endothelial injury and repair in childhood cancer patients treated with anthracyclines by quantifying circulating endothelial cells (CECs) and endothelial progenitor cells (EPCs).

**Methods:** In a single-centre retrospective cohort, children (<18 years) diagnosed with leukemia (n=35) or lymphoma (n=13) were studied at four timepoints: pre-treatment (“Pre”), ∼1-month- (“End”), 3 months- (3M), and 1 year- (1Y) post-treatment. Peripheral blood mononuclear cells were analyzed by flow cytometry to quantify CECs and EPCs, and EPC fate was assessed by p16 (senescence) and Annexin V (apoptosis). Cardiac injury biomarkers and left ventricular function were assessed at each timepoint.

**Results:** Longitudinal trends of CEC and EPC counts were similar between leukemia and lymphoma participants. CECs were highest at pre-treatment and declined significantly thereafter, though they remained marginally elevated during remission compared with healthy controls, indicating that endothelial damage had largely subsided following treatment. EPCs were also highest at pre-treatment and decreased to levels below healthy controls during remission, suggesting impaired baseline endothelial maintenance and repair. Furthermore, EPCs were predominantly senescent up to 1-year post-treatment.

**Conclusions:** Endothelial injury resolves by treatment completion, but repair remains impaired during remission with EPC pools dominated by senescent cells. This suggests defective endothelial regeneration, rather than persistent injury, drives long-term cardiovascular complications and underscores the need to restore EPC viability and function in childhood cancer survivors.

## 1. Introduction

Anthracycline is a class of chemotherapeutic agents used to treat more than half of all childhood cancers^1^. Despite their effectiveness in treating cancers, anthracyclines are associated with inducing cytotoxicity in other cell types resulting in adverse outcomes^2^. For example, cardiotoxicity represents a particularly prevalent complication where up to 57% of children who have been treated with anthracyclines exhibit cardiac abnormalities^3^, and up to 16% develop congestive heart failure^4^. Additionally, late-onset cardiotoxicity which occur years after treatment completion, is exhibited via impaired myocardial growth and function^4,5^.

Given that endothelial cells lining the blood vessels serve as a protective barrier for cardiac cells and are the most abundant cell type in the heart, their impairment and cell death can exacerbate acute and late-onset anthracycline-mediated cardiac injury^6–11^. It is well-established that endothelial dysfunction increases the risk of cardiovascular complications^12^. Thus, to assess endothelial health, we enumerated circulating endothelial cells (CECs) and endothelial progenitor cells (EPCs) in the peripheral circulation of pediatric leukemia and lymphoma patients who were treated with anthracyclines. CEC is a putative marker of endothelial injury, while EPC is regarded as marker of endothelial repair and neovascularization ^13–15^. In addition, cardiac injury markers present in plasma such troponin T (TnT) and N-terminal pro B-type natriuretic peptide (NT-proBNP) were assayed to elucidate the potential impact of endothelial health on anthracycline-mediated cardiac injury.

Leukemia is a hematological malignancy which begins in the bone marrow where hematopoietic cells proliferate aberrantly into immature forms, functionally impairing bone marrow^16^. The uncontrolled proliferation exacerbates hypoxia in the bone marrow triggering hypoxic inducible factors which stimulate angiogenesis and mobilize EPCs into the circulation^17–19^. Furthermore, systemic inflammation in leukemic patients can trigger the mobilization of EPCs from the bone marrow to the site of tissue injury^18,20^. Other potential sources of circulating EPCs include vascular resident endothelial cells and tissue vascular niches^21^. In the setting of cancer, EPCs are indicative of either vascular repair or severity of cancer and need to be interpreted with caution^22^. Separately, high CEC counts have been reported in adult leukemia patients when compared to treated patients and healthy controls^23,24^, potentially due to systemic inflammation^25,26^ or the disruption of existing vasculature (generating CECs) facilitating angiogenesis in the bone marrow^19,27^. However, to date, no studies have longitudinally monitored CEC and EPC counts in childhood leukemia patients up to one-year post-treatment, a period during which early-onset cardiotoxicity often develops^28^.

Lymphoma develops in the lymphatic system^29^. It emerges due to genetic mutation in a lymphocyte causing clonal expansion which results in accumulation of cells in the lymph node^29^. The progression of lymphoma is enabled by vascularization, also known as “angiogenic switch”, which promotes the delivery of oxygen and nutrients necessary for cancer cell survival and proliferation^30,31^. Lymphoma cells release pro-angiogenic growth factors into the cancer microenvironment and recruit EPCs to the angiogenic site which either differentiate into endothelial cells or release angiogenic growth factors for blood vessel formation^31–33^. The inflammatory-laden setting and disruption of blood vessels to facilitate angiogenesis can cause CECs to be elevated. As expected, the amount of EPCs and CECs are higher in adult lymphoma patients when compared to healthy controls ^34,35^. However, there are no reports of CEC and EPC counts on acute childhood lymphoma.

In addition to enumerating EPCs at different timepoints, the proportion of EPCs committed to the senescent and apoptotic fates were characterized in both leukemia and lymphoma patients. Emerging evidence indicates that anthracyclines trigger both apoptotic and senescent pathways in EPCs, both of which impair EPC function^36^. EPC fates will shed light on their capability on maintaining vascular health. While the effect of EPC cell fate on acute- and late-onset cardiotoxicity has not been explored before, the restoration of EPC function ameliorated doxorubicin-induced cardiac dysfunction in rat model^9^. Thus, we characterized EPC cell fates, specifically apoptotic and senescent EPCs, in anthracycline-treated pediatric patients.

This study is the first to report longitudinal trends in CECs, EPCs, and cardiomyocyte injury biomarkers in childhood cancer patients from the time of diagnosis to one-year post-anthracycline treatment. Additionally, the cell fate of EPCs was characterized to assess its usefulness in facilitating endothelial health. Having a comprehensive understanding of the course of endothelial health and injury will shed light on the initial asymptomatic insult leading to late-onset cardiovascular complications^11,37^. These markers have the potential to guide diagnosis and therapeutic decision-making processes and predict prognosis^38^. These efforts are necessary with the increasing numbers of childhood cancer survivors^39^.

## 2. Methodology

### 2.1 Study Population

This was a single-centre, retrospective cohort study that was approved by the Domain Specific Review Board of National Healthcare Group, Singapore. Children (<18 years old) diagnosed with cancer who are due to receive anthracyclines as part of combination chemotherapy were recruited between 2019 and 2024 at National University Hospital (NUH). In addition, only children with normal cardiac function prior to initiation of anthracycline therapy (LVEF > 55%) were included in the study. Clinical parameters were obtained from EPIC, an electronic health record system used in NUH. Peripheral blood and echocardiograms were obtained at four timepoints for this study: before anthracycline treatment (Pre), one month after treatment (End), three months after treatment (3M) and one year after treatment (1Y).

### 2.2 Blood Processing

Approximately 10mL of blood was collected in EDTA tubes at each timepoint and centrifuged at 4000rpm for 10 min at 4°C within 2 hours of collection. Plasma was transferred to a clean tube and gently resuspended to get a homogenous mixture before transferring to cryotubes for storage. Subsequently, buffy coats were transferred into cryotubes. Both plasma and buffy coats were stored at -80°C until analysis.

### 2.3 Circulating Endothelial Cell and Endothelial Progenitor Cell Enumeration Via Flow Cytometry

To recover mononuclear cells, frozen buffy coats were thawed and diluted threefold with phosphate-buffered saline (PBS). The diluted buffy coat was layered over half volume of Ficoll-Paque™ PLUS density gradient media and centrifuged at 1000g for 20 minutes with an acceleration setting at 1 and no deceleration, at room temperature. The layer of mononuclear cells was transferred to a new tube, mixed with an equal volume of PBS and subsequently centrifuged at 540g for 7 minutes at maximum acceleration and deceleration. The resulting cell pellet was washed with 5mL of PBS and centrifuged at the same setting. The cell pellet was resuspended in 500µL of 1% bovine serum albumin (BSA) in PBS for cell counting. Aliquots of 1.5 million cells were distributed into individual tubes for antibody staining and remaining cells were cryopreserved for future use. Each tube of cells was stained with 4 antibodies: CD45 (FITC), CD31 (PE-Cy7), CD133 (APC) and Hoechst 33342. Following a 10-minute incubation at room temperature, the cells were incubated in the cold room (∼4°C) on a see-saw rocker for 20 minutes. After incubation, 1 mL of 1% BSA in PBS was added to the cells to dilute excess antibodies, followed by centrifugation at 300g for 5 minutes at 4°C to remove them. The resulting cell pellet was resuspended in 200µL of 1% BSA in PBS and filtered through a 60µm filter to remove clumps, yielding a single-cell suspension. The cell suspension was analyzed using the Beckman Coulter CytoFLEX LX Flow Cytometer, with 10000 cells analyzed per tube. CECs were identified based on the immunophenotypic profile CD45^−^/CD31^+^/CD133^−^ /DNA^+^, while EPCs were identified by CD45^−^/CD31^+^/CD133^+^/DNA^+^.

### 2.4 Endothelial Progenitor Cell Fate Characterization Via Flow Cytometry

Using the remaining cryopreserved cells, EPCs were further characterized for apoptosis using Annexin V staining and senescence using p16 staining. Annexin V antibody was conjugated to Pacific Blue fluorophore with an emission wavelength overlapping that of Hoechst 33342. Therefore, Hoechst staining was excluded during the characterization of apoptotic EPCs, which were identified using the immunophenotypic profile CD45⁻/CD31⁺/CD133⁺/Annexin V⁺. Annexin V binds to phosphatidylserine on the cell surface in a calcium-dependent manner, necessitating the use of a calcium-containing binding buffer. The cells were first stained with CD45, CD31 and CD133 antibodies as described in section 2.3. Following the final centrifugation, the resulting cell pellet was resuspended in 100µL of binding buffer (BioLegend #422201) followed by addition of Annexin V antibodies. The cells were incubated for 15 minutes at room temperature in the dark to facilitate binding. Finally, 200 µL of binding buffer was added to achieve an optimal volume of 300 µL for flow cytometric analysis. p16 antibody was conjugated to CoraLite^®^555. Senescent EPCs were identified by an immunophenotypic profile of CD45^−^/CD31^+^/CD133^+^/p16^+^. To achieve this, the cells were first stained with all antibodies, except p16, as described above. Prior to staining with p16 antibody, the cells were fixed by incubating the cells in 500µL of fixation buffer (BioLegend #420801) for 20 minutes at room temperature. After which the cells were centrifuged at 350g for 5 minutes and subsequently resuspended in 500 µL of 1× permeabilization and wash buffer (BioLegend #421002). This permeabilisation step was repeated once more. After the final centrifugation, most of the supernatant was removed, leaving 100 µL of residual buffer. Cells were resuspended in the residual buffer and 1µL of p16 antibody was added. The cells were incubated for 20 minutes at room temperature in the dark. Following incubation, 1 mL of 1× permeabilization and wash buffer was added to dilute unbound antibodies, and the cells were centrifuged at 350g for 5 minutes. The pellet was resuspended in 300 µL of 1× permeabilization and wash buffer and filtered through a 60 µm filter to remove cell clumps. The single-cell suspensions were analyzed using the Beckman Coulter CytoFLEX LX Flow Cytometer, with 10000 cells analyzed per tube.

### 2.5 Echocardiography

All echocardiographic studies will be performed using the Lisendo 880 System (Hitachi). All offline measurements will be performed using the Tomtec vendor-independent analysis software (Tomtec, USA). Left ventricular ejection fraction was derived from echocardiographic measurements to assess cardiac function throughout the study period.

### 2.6 Cardiac Protein Assay

High-sensitivity cardiac troponin T (hsTnT) and N-terminal pro B-type natriuretic peptide (NTproBNP) proteins were assayed with Roche Diagnostics, Cobas e411 automated analyzer using 300µL of plasma from the respective timepoints. TnT is released into the peripheral circulation upon irreversible cardiomyocyte injury, thus becoming a robust marker of cardiomyocyte damage^40^. NTproBNP is synthesized and secreted by ventricular cardiomyocytes in response to increased ventricular wall stress making it a robust marker of heart failure and cardiac dysfunction^41^.

### 2.7 Statistical Analysis

Statistics were calculated using GraphPad Prism 7.2 (GraphPad Software, San Diego, CA, USA). The normality of the sample populations was determined using Kolmogorov-Smirnov test and homogeneity was determined using F-test, prior to conducting Student’s t-test. One-tailed Student’s t-tests were performed to determine statistical significance between the means of two groups. If the sample populations were nonparametric, Mann–Whitney test was employed. If they were parametric but not homogenous, Student’s t-test with Welch’s correction was applied. All data are shown as means ± S.E.M. Spearman correlation was employed as Kolmogorov-Smirnov test determined populations to be nonparametric.

## 3. Results

### 3.1 Demographics and Clinical Parameters of Patients

35 patients diagnosed with leukemia were recruited. Among these, 68% had acute lymphoblastic leukemia (ALL), 23% had acute myeloid leukemia (AML) and the remaining cases were neonatal acute or B-cell acute lymphoblastic leukemia. Separately, 13 patients diagnosed with lymphoma were recruited. 38% were diagnosed with Hodgkin lymphoma, 38% with Burkitt lymphoma, and the remainder with anaplastic large-cell, B-cell or primary mediastinal B-cell lymphoma. Age-matched comparisons were performed for CEC and EPC enumeration experiments. Anthracycline cumulative dose and left ventricular ejection fraction (%LVEF) are similar in both leukemia and lymphoma participants across all time points of the study (Table 1). It has been confirmed by a team of oncologists that these research participants are in remission by end-timepoint, based on leukemic blast counts and PET-CT data.

**Table 1:** Demographics of research participants.

|  | **Leukemia**<br>**(n=35)** | **Lymphoma**<br>**(n=13)** | **Healthy**<br>**(n=12)** | **p-value** |
| --- | --- | --- | --- | --- |
| **Female (n)** | 11 | 3 | 3 | - |
| **Male (n)** | 24 | 10 | 9 | - |
| **Participants' age for CEC and EPC enumeration (years ± SEM)** | 8.14 ± 0.96 | - | 10.7 ± 1.7 | 0.1127 |
| **Participants' age for CEC and EPC enumeration** | - | 12.6 ± 0.83 | 11.5 ± 1.65 | 0.3409 |
| **Participants' age for EPC characterization** | 8.3 ± 1.1 | - | 15.1 ± 1.2 | 0.0012 |
| **Participants' age for EPC characterization** | - | 12.6 ± 0.9 | 15.1 ± 1.2 | 0.04 |
| **Cumulative anthracycline dose (mg/m<sup>2</sup>)** | 177 ± 15.4 | 182.3 ± 24.3 | 0 | 0.25 |
| **%LVEF** | 61.4 ± 0.76 | 60.9 ± 0.7 | - | 0.23 |

Separately, neutrophil-to-lymphocyte ratio (NLR) was used to assess systemic inflammation. Although the threshold for childhood cancers is not established^42,43^, typically, NLR>3 reflects systemic inflammation. Based on this, pronounced inflammation was observed at 3 months post-treatment (NLR = 5.7) in participants with leukemia and at pre-treatment (NLR = 7.4) in those with lymphoma (Table 2).

**Table 2:**
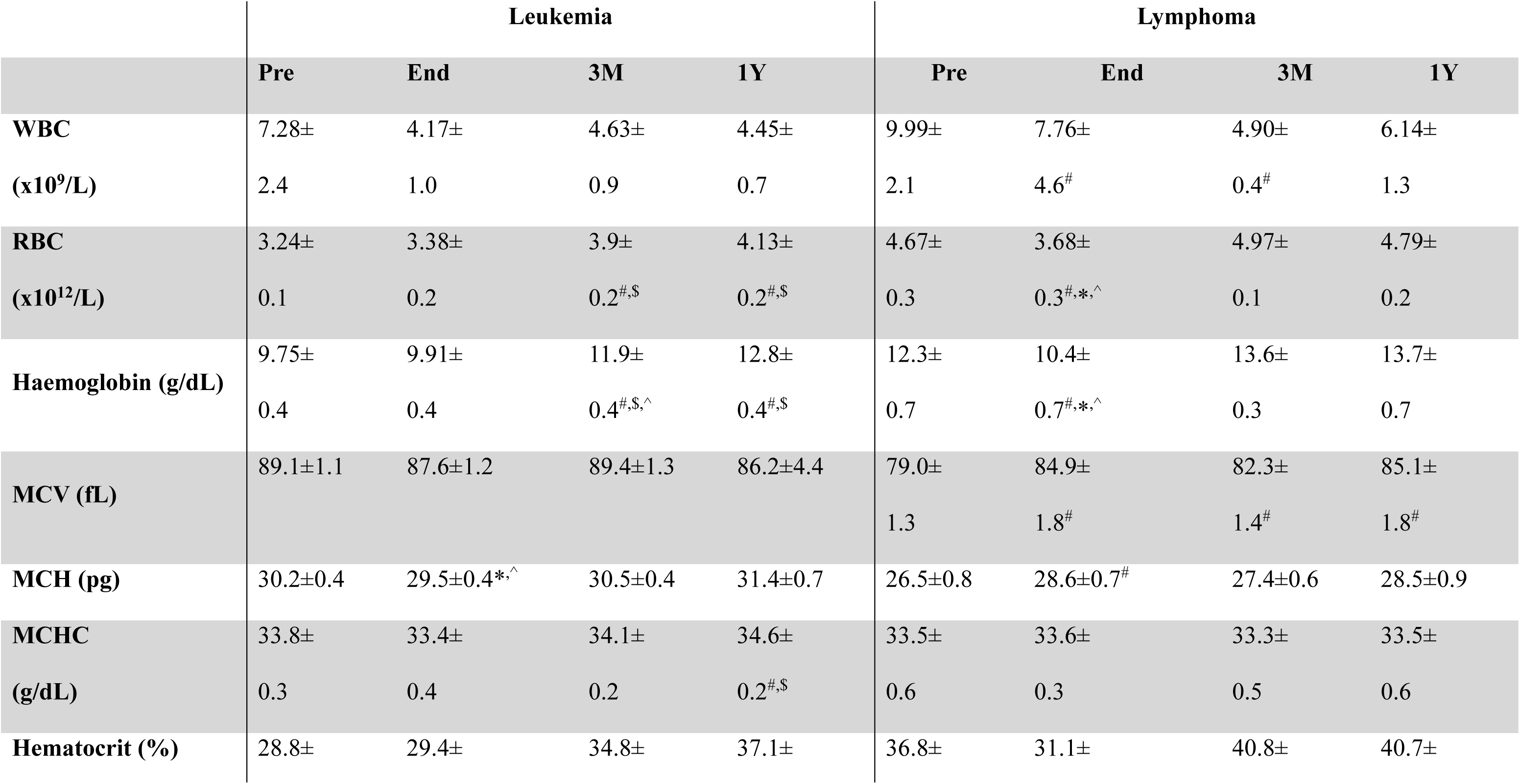
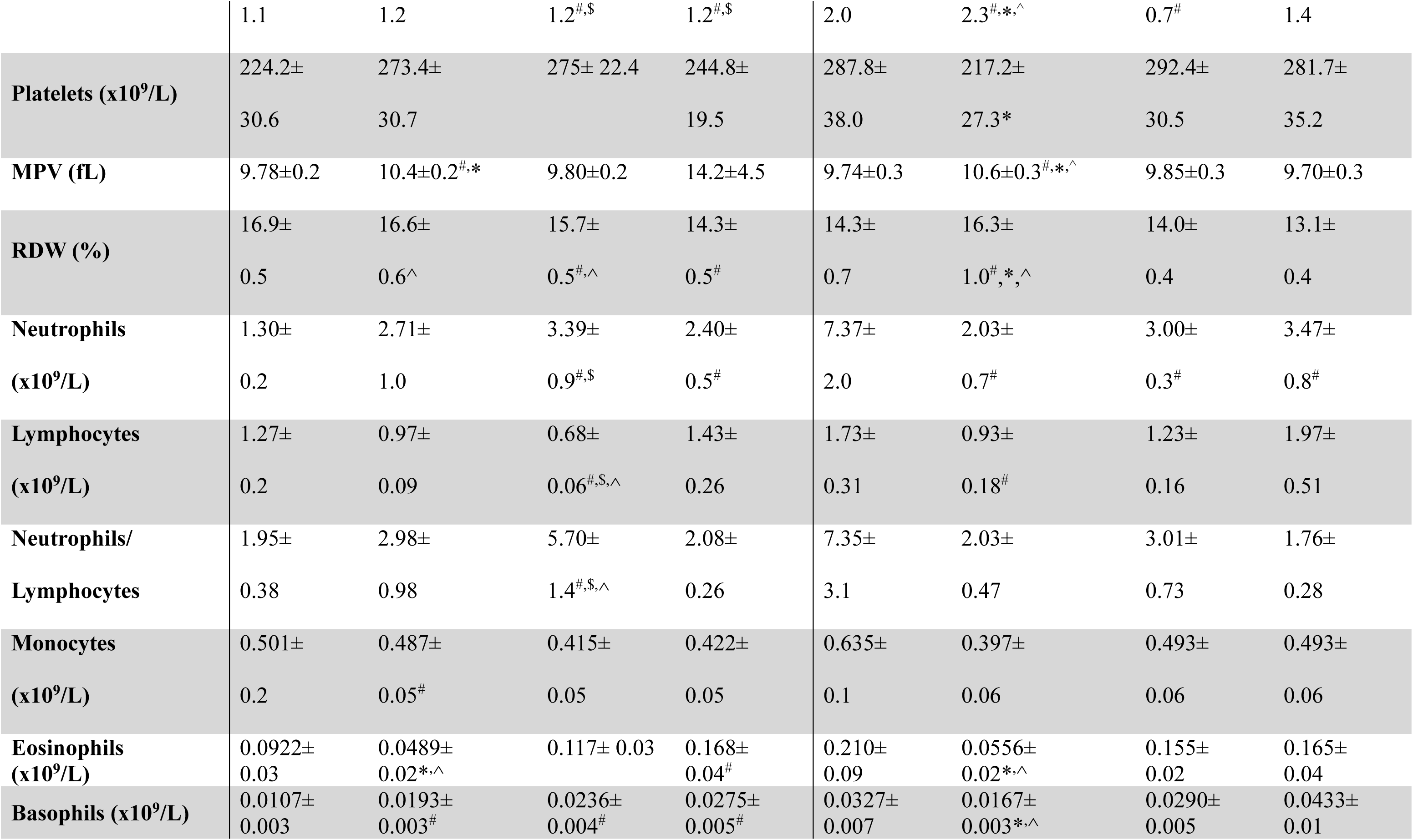
Clinical laboratory characteristics of research participants. #, $, * and ^ symbols indicate statistically significant differences with Pre, End, 3M and 1Y timepoints, respectively.

### 3.2 Peripheral CEC and EPC Enumeration in Leukemia Patients

CEC were in the range between 0.35% and 1.2% of 10,000 peripheral blood mononuclear cells (PBMCs). Levels were highest at the time of diagnosis (i.e., pre-treatment) where CEC counts were approximately 4 folds higher than healthy controls. CECs then decreased after treatment (i.e., End-, 3M- and 1Y-timepoints) to levels comparable to healthy controls, although remaining marginally higher (Figure 1B&C). This indicates that the processes of cancer itself cause more endothelial damage than the residual effects of anthracycline treatment. The circulating EPC count is between 0.12% and 2.6% of 10,000 PBMCs. EPC counts at the pre-treatment is comparable to healthy controls. However, the counts significantly decreased at End, 3M and 1Y timepoints when compared to healthy controls, implying that endothelial repair is impaired after treatment (Figure 1D&E). Collectively, these findings suggest that persistent endothelial impairment during remission is more likely attributable to defective repair mechanisms than to active residual chemotherapy-induced injury.

**Figure 1:**
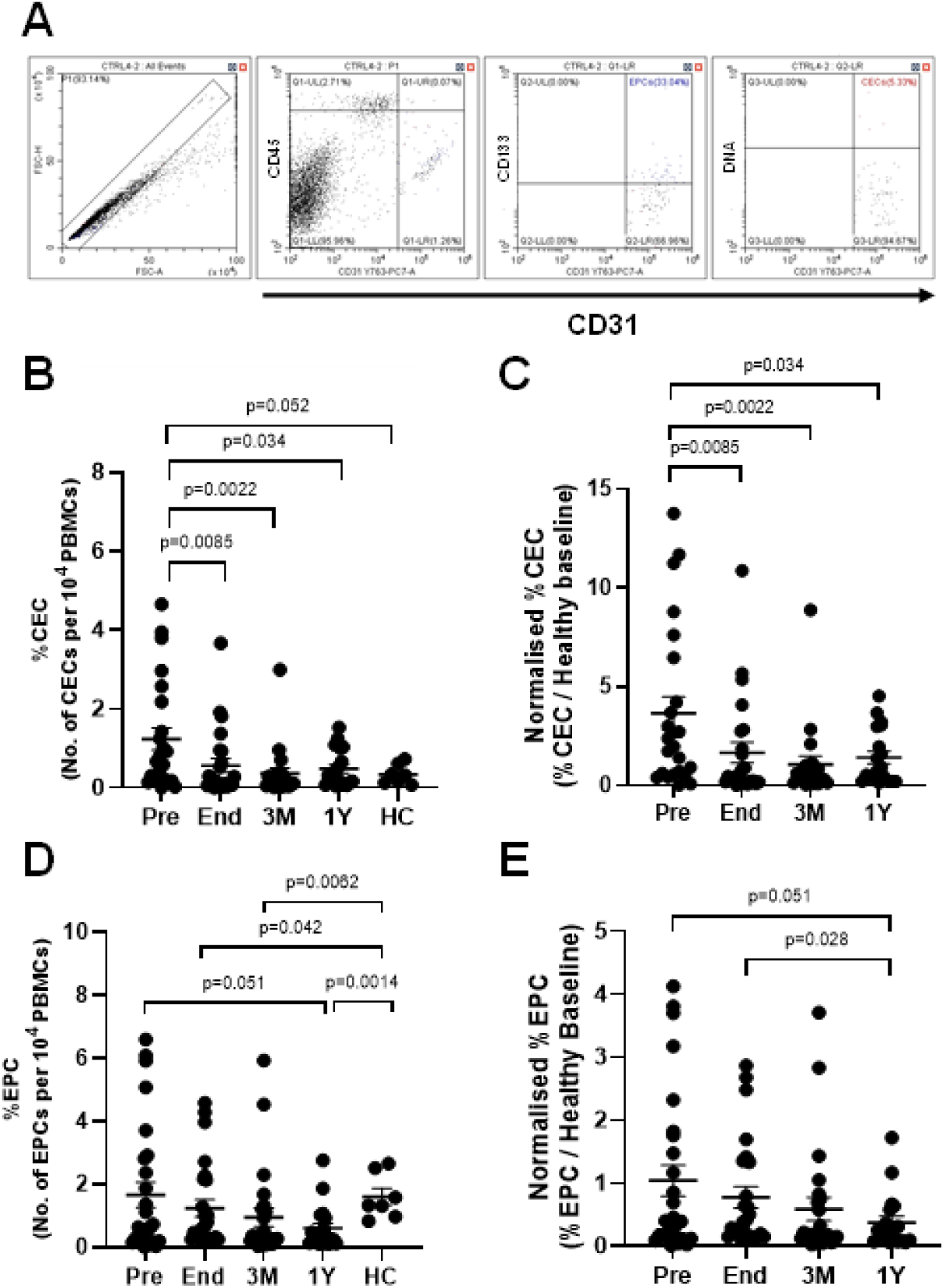
Enumeration of CECs in Leukaemic patients at Pre-treatment (n=24), End-treatment (n=24), 3 months post-treatment (n=21), 1 year post-treatment (n=19) and healthy controls (n=7). **(A)** Gating strategy employed on flow cytometry result to identify CECs and EPCs **(B)** %CECs in 10,000 PBMCs. **(C)** %CECs at each timepoint are normalized by %CEC in healthy controls. **Enumeration of EPCs in Leukemic patients at Pre-treatment (n=27), End-treatment (n=25), 3 months post-treatment (n=24), 1 year post-treatment (n=19) and healthy controls (n=7). (D)** %EPCs in 10,000 PBMCs. **(E)** %EPCs at each timepoint are normalized by %EPC in healthy controls. Data is presented as a scatterplot of mean ± SEM. Numerical values are presented in the supplementary materials (Table S1). Statistical tests employed are described in the methodology section.

### 3.3 Endothelial Progenitor Cell Fate Characterization in Leukemia patients

Figure 1D depicts that EPCs are decreased in the peripheral blood circulation between End and 1Y timepoints. While this in itself contributes to disrupted endothelial repair, it is important to understand the viability of the available cells to comprehensively evaluate their reparative capacity. This is because the cytotoxic effects of anthracyclines trigger apoptotic and senescent pathways in cells^44,45^. Thus, we examined the fraction of EPCs that have committed to the senescent or apoptotic pathway via flow cytometry.

In this study, the senescent p16^+^ EPCs were in the range between 73 and 85% of all EPCs detected (Figure 2C). p16⁺ EPCs were elevated in leukemic participants at all timepoints relative to healthy controls, particularly at the end-timepoint.

**Figure 2:**
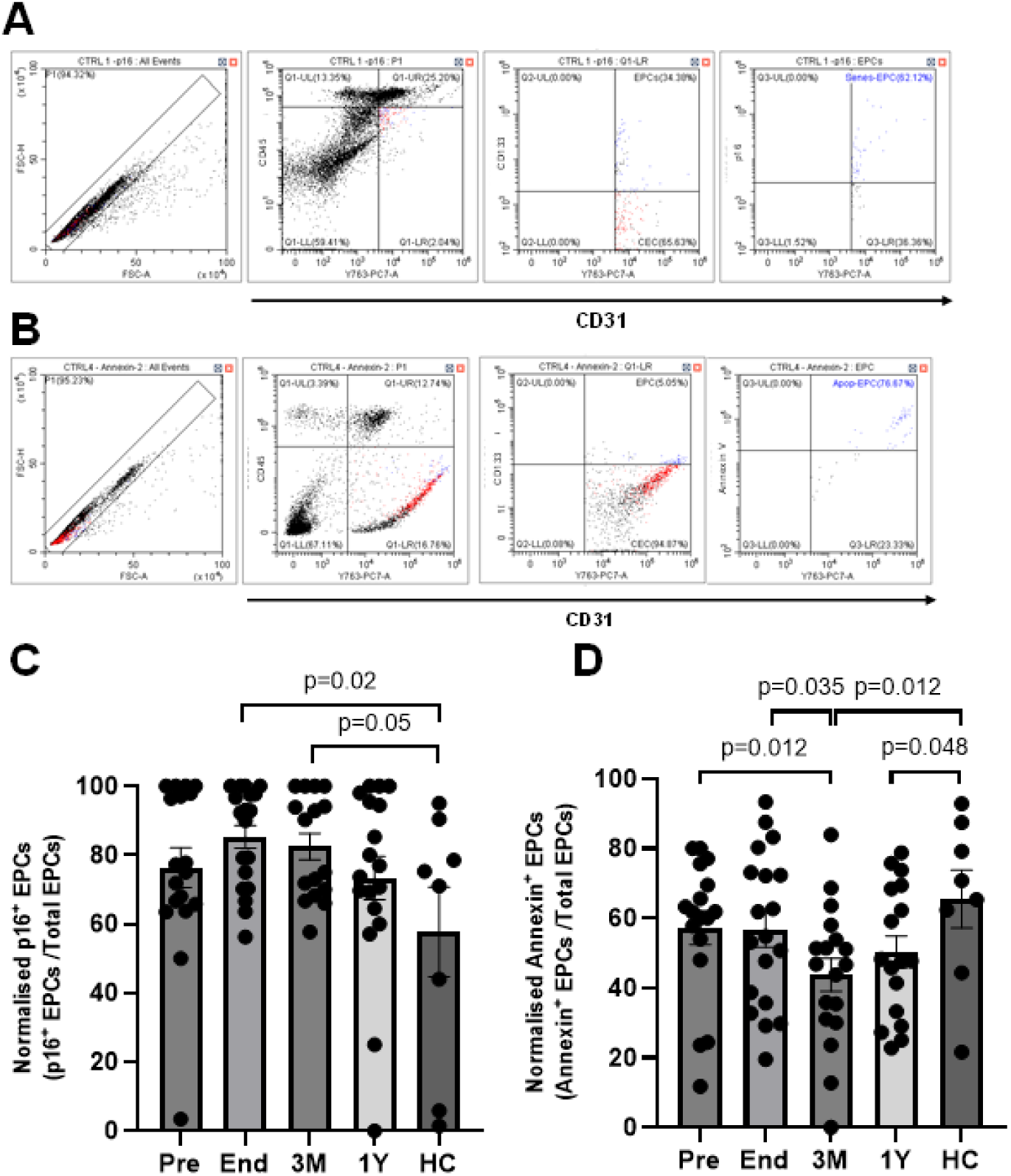
EPC characterisation in Leukemic patients at Pre-treatment (n=18), End-treatment (n=19), 3 months post-treatment (n=16), 1 year post-treatment (n=18) and healthy control (n=8). **(A)** Gating strategy employed on flow cytometry data to identify senescent EPCs **(B)** Gating strategy employed on flow cytometry data to identify apoptotic EPCs **(C)** Percentage of EPCs positive for p16 signal relative to total EPCs **(D)** Percentage of EPCs with positive Annexin V signal relative to total EPCs. Data are presented as a scatterplot of mean ± SEM. Statistical tests employed are described in the methodology section.

On the other hand, the apoptotic Annexin V^+^ cells were in the range between 44 and 57% of all EPCs detected. Unlike senescent EPCs, the proportion of apoptotic EPCs at all time points were lower than healthy controls. The data shows that there are relatively higher apoptotic EPCs at pre- and end-treatment timepoint compared to 3 months post-treatment (Figure 2D). The significant decrease in apoptotic cells at 3 months post-treatment could be attributed to the nature of anthracycline chemotherapy as doxorubicin-induced apoptosis is reported to occur acutely^46^. Thus, the associated apoptotic effects are potentially subsiding by 3 months post-treatment. Additionally, the reduction in apoptotic EPCs may result from increased cellular senescence particularly at 3 months post-treatment, resulting in apoptotic EPCs to decrease below healthy baseline, as senescent cells often upregulate anti-apoptotic pathways that counteract apoptosis^47^. Altogether, these data demonstrates that a significant proportion of circulating EPCs develop senescence after treatment, reiterating that any endothelial impairment during remission is likely due to defective repair mechanisms.

### 3.4 Cardiac Protein Biomarkers in Leukemia Patients

Majority of the patients had hsTnT levels below the marker of elevation before treatment (7.34±1.6 pg/mL). However, the hsTnT significantly increased at end-treatment (16.2±2.3pg/mL), indicating significant increase in cardiomyocyte damage. The levels significantly decreased from end- to 3 months post-treatment (11.0±1.5 pg/mL) with 37% of the participants remaining above the marker of elevation. By 1-year post-treatment, the hsTnT levels of almost all participants returned to acceptable levels (5.53±0.74) (Figure 3A). On the other hand, NTproBNP levels of the participants were mostly in the acceptable range throughout the study period implying that there is no significant cardiac remodeling despite cardiomyocyte damage (Figure 3B). This finding is further supported by the preservation of %LVEF within the normal range of these participants. Spearman test was performed to identify the correlation between hsTnT (cardiomyocyte damage) and CECs (endothelial damage) and hsTnT and EPCs, respectively. However, there were no significant correlations revealed (Figure S1). The main limitation of this approach lies in the non–organ-specific nature of CECs. While higher CEC levels suggest increased endothelial shedding, it cannot be definitively concluded that these cells originate from the coronary vasculature, thereby limiting the accuracy of linking cardiac endothelial injury to cardiomyocyte damage. Hence, using organ-specific EC injury markers like cell-free DNA will be useful to elucidate whether damage to cardiac- or coronary-specific endothelium exacerbates cardiomyocyte damage or vice versa.

**Figure 3:**
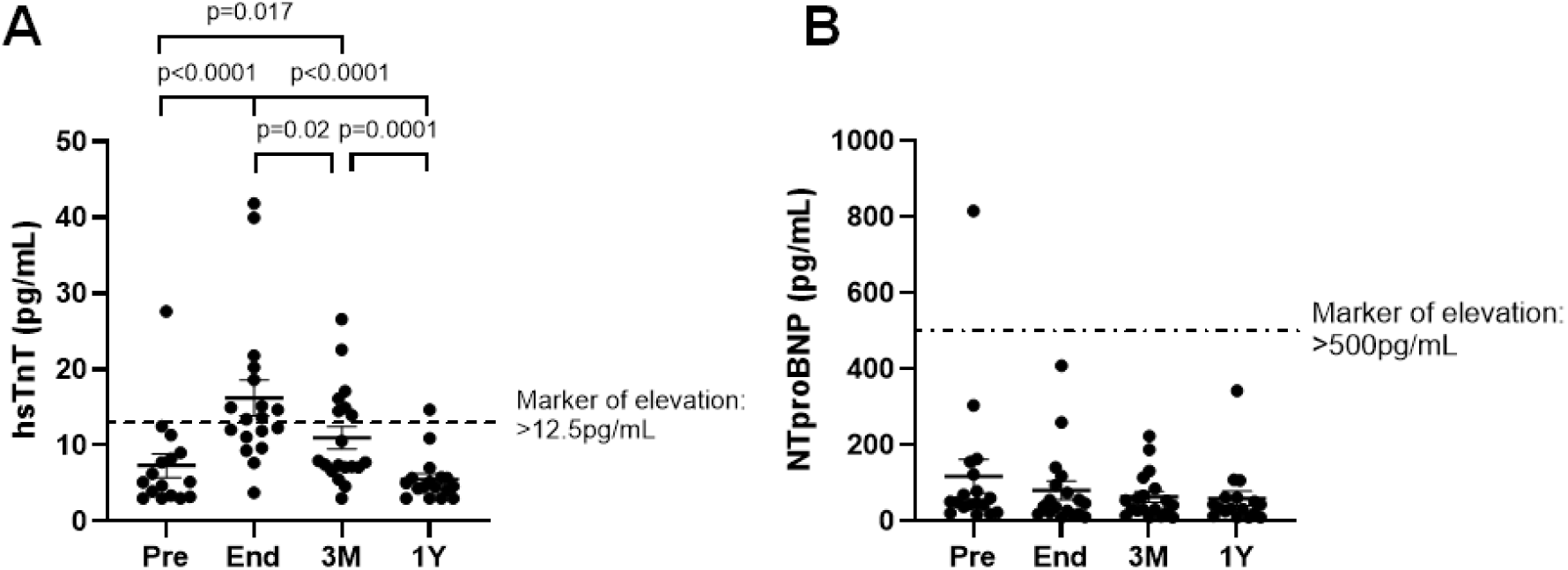
Assay of cardiac biomarkers in Leukemic patients at Pre-treatment (n=16), End-treatment (n=18), 3 months post-treatment (n=19), 1 year post-treatment (n=17). **(A)** Assay of High-sensitivity troponin T (hsTnT) **(B)** Assay of NTproBNP. Data is presented as a scatterplot of mean ± SEM. Statistical tests employed are described in the methodology section.

### 3.5 Peripheral CEC and EPC Enumeration in Lymphoma Patients

CECs were in the range between 0.25% and 1.74% of 10,000 PBMCs. Just as in leukemia participants, lymphoma patients exhibited the highest CEC levels at diagnosis, with counts approximately 5.5-fold higher than those of healthy controls (Figure 4A&B). This elevation is consistent with the notion that endothelial disruption is required to support angiogenesis during lymphoma progression. Additionally, depending on the cancer severity, inflammatory-laden environment may exacerbate vascular injury, contributing to CECs^48^. Enhanced inflammation at pre-treatment is reflected by the NLR (Table 2). Although CEC levels decreased to values comparable to healthy controls at the end of treatment, they were approximately 2-folds higher at 3 months and 1-year post-treatment compared to healthy controls, albeit without statistical significance (Figure 4A). These suggest that persistent endothelial impairment is attributable to residual anthracycline-induced endothelial injury. Due to the nature of lymphoma, circulating EPCs may reflect not only endothelial repair but also cancer-driven angiogenesis, and thus serve as an indicator of cancer progression. The circulating EPC ranges between 0.26% and 2.6% of 10,000 PBMCs (Figure 4C&D). However, the counts are significantly decreased at End, 3M and 1Y timepoints when compared to pre-treatment and healthy controls, respectively. The significant decrease indicates that the progression of lymphoma is impeded, which corroborates with the absence of significant masses in PET-CT scans. However, a significant decrease compared to healthy controls indicates that basal endothelial repair is impaired. Collectively, the CEC and EPC profiles suggest that persistent endothelial impairment during remission is driven predominantly by reduced reparative capacity, with a lesser contribution from residual anthracycline-induced vascular injury.

**Figure 4:**
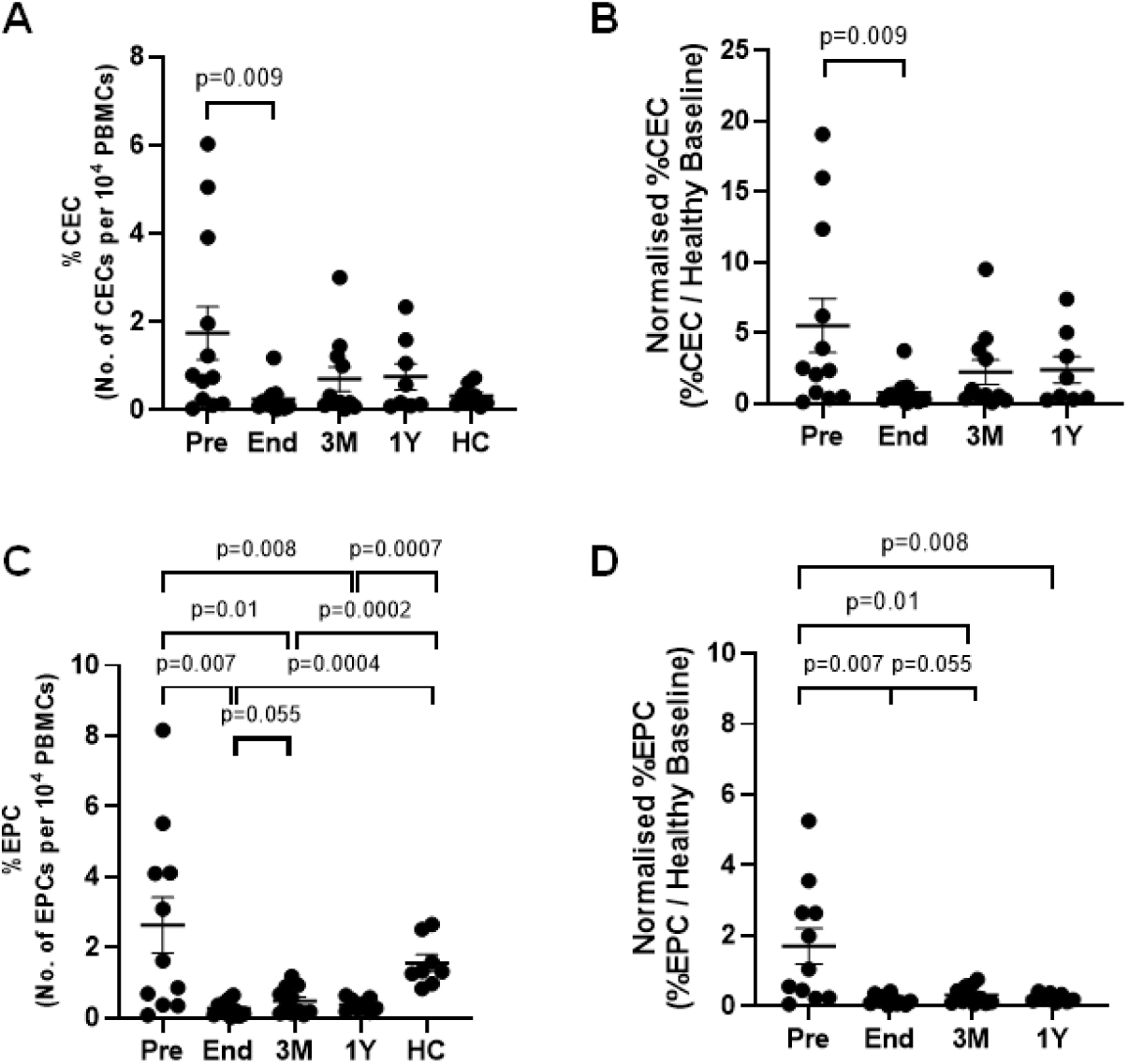
Enumeration of CECs in Lymphoma patients at Pre-treatment (n=12), End-treatment (n=11), 3 months post-treatment (n=11), 1 year post-treatment (n=8) and healthy controls (n=8). **(A)** %CECs in 10,000 PBMCs. **(B)** %CECs at each timepoint are normalized by %CEC in healthy controls. **Enumeration of EPCs in Lymphoma patients at Pre-treatment (n=11), End-treatment (n=11), 3 months post-treatment (n=11), 1 year post-treatment (n=8) and healthy controls (n=8). (C)** %EPCs in 10,000 PBMCs. **(D)** %EPCs at each timepoint are normalized by %EPC in healthy controls. Data is presented as a scatterplot of mean ± SEM. Numerical values are presented in the supplementary materials (Table S2). Statistical tests employed are described in the methodology section.

### 3.6 Endothelial Progenitor Cell Fate Characterization in Lymphoma patients

In this study, the senescent p16^+^ EPCs are generally in the range between 73 and 95% of all the EPCs detected. Just as in leukemia patients, p16⁺ EPCs were significantly elevated in lymphoma participants at all time points relative to healthy controls, indicating that the cancer environment and cytotoxic nature of anthracyclines could have triggered senescence pathway in EPCs (Figure 5A). The apoptotic Annexin V^+^ cells were in the range between 23 and 62% of all the EPCs detected. Unlike senescent EPCs, the proportion of apoptotic EPCs were lower than in healthy controls at all time points. This study shows that apoptotic EPCs are significantly lower at pre-treatment compared to healthy controls (Figure 5B). Although this is not established, this observation is expected as the EPCs are required to vascularize and support lymphoma tumors^49^. However, it continued to increase up to 1-year post-treatment, although it is still marginally lower than healthy controls. Again, the data demonstrates that the proportion of apoptosis increases as senescence decreases, reiterating that senescence opposes apoptosis. Altogether, this data suggests that senescent EPC is predominant and emphasizes that endothelial impairment during remission is likely due to defective repair mechanisms.

**Figure 5:**
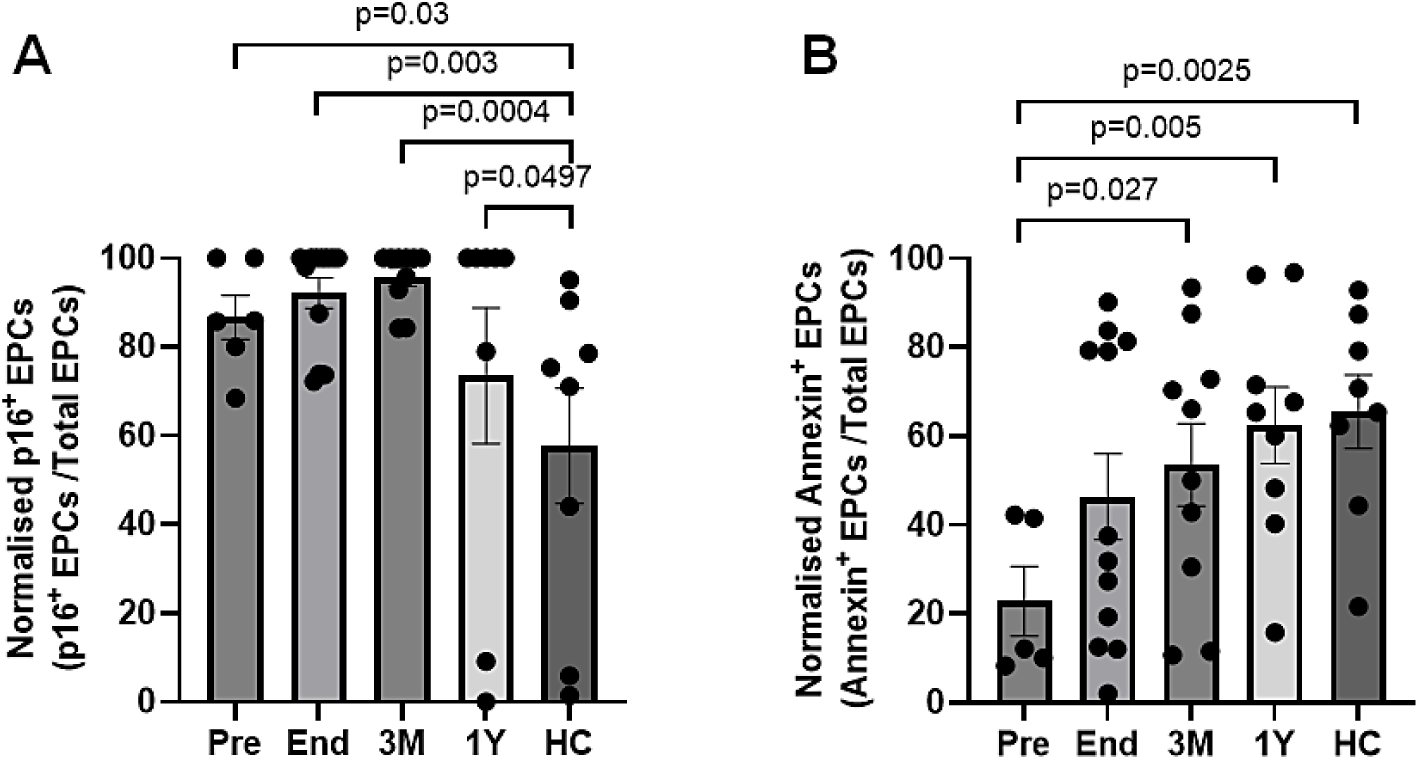
EPC characterization in Lymphoma patients at Pre-treatment (n=6), End-treatment (n=12), 3 months post-treatment (n=10), 1 year post-treatment (n=8) and healthy control (n=8). **(A)** Percentage of EPCs positive for p16 signal relative to total EPCs **(B)** Percentage of EPCs with positive Annexin V signal relative to total EPCs. Data is presented as a scatterplot of mean ± SEM. Statistical tests employed are described in the methodology section.

### 3.7 Cardiac protein biomarkers in Lymphoma patients

All patients had hsTnT levels below the marker of elevation before treatment (4.45±0.4 pg/mL). Although hsTnT significantly increased at end-treatment (11.5±2.7 pg/mL) indicating significant cardiomyocyte damage, only two participants had levels above the marker of elevation. The levels decreased from end- to 3 months post-treatment (8.88±1.5 pg/mL) with all except one participant returning to below the marker of elevation. By 1-year post-treatment, the hsTnT levels of almost all participants returned to acceptable levels (5.67±1.7 pg/mL) (Figure 6A). Collectively, the data suggests marginal cardiomyocyte damage occurred in lymphoma participants at end-timepoint. Additionally, NTproBNP levels of all participants after treatment were in the acceptable range implying that there is no significant cardiac remodeling (Figure 6B). This finding is supported by the preservation of %LVEF within the normal range. Spearman test was performed to identify the correlation between hsTnT (cardiomyocyte damage) and CECs (endothelial damage) and hsTnT and EPCs, respectively. However, there were no significant correlations revealed (Figure S2). This could be due to the lack of significant cardiomyocyte damage in the lymphoma population of this study.

**Figure 6:**
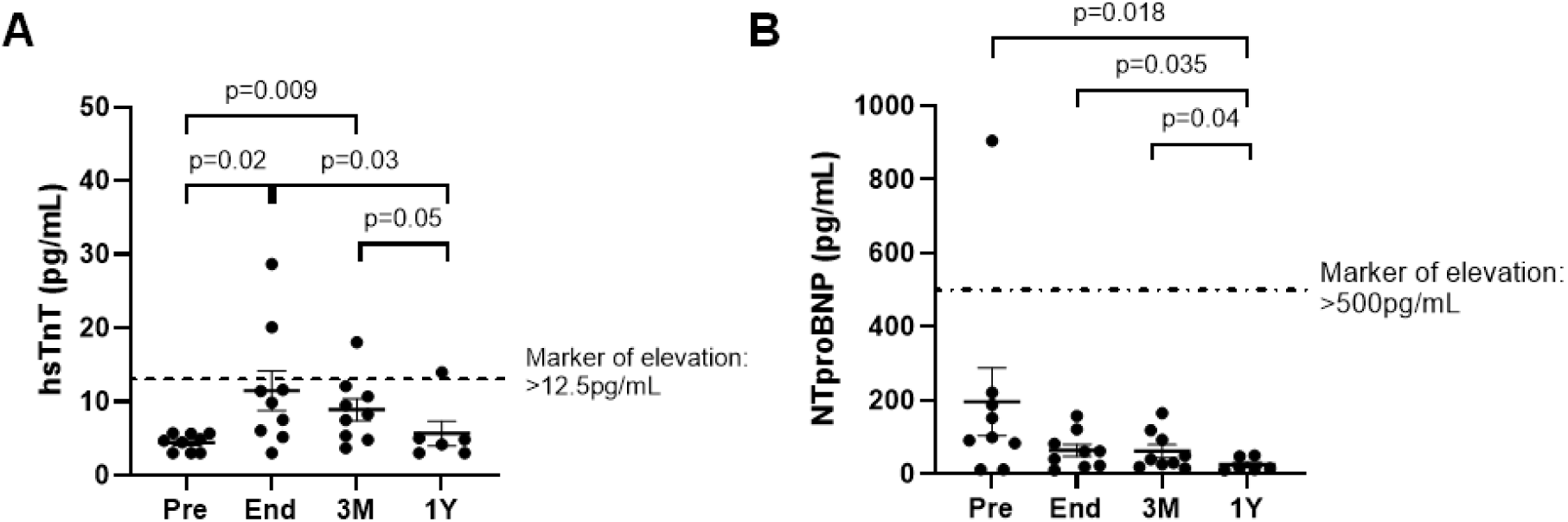
Assay of cardiac biomarkers in Lymphoma patients at Pre-treatment (n=9), End-treatment (n=9), 3 months post-treatment (n=9), 1-year post-treatment (n=6). **(A)** Assay of High-sensitivity troponin T (hsTnT) **(B)** Assay of NTproBNP. Data is presented as a scatterplot of mean ± SEM. Statistical tests employed are described in the methodology section.

## 4. Discussion

It is established that anthracyclines represent a double-edged sword. That is while they are highly effective anticancer agents, their use is limited by the risk of acute and chronic cardiotoxicity. Since the 1970s, research on anthracycline-induced cardiotoxicity has predominantly focused on cardiomyocytes. Given that the muscle to capillary ratio is 1:1^6,50^ and the proximity between capillary endothelial cells and cardiomyocytes (i.e., 1µm), it is realistic that anthracycline-related cardiac dysfunction is caused not only by impaired cardiomyocytes but also facilitated by compromised endothelial cells^6^. Hence, over the past decade, increasing attention has been directed toward the role of the vascular endothelium in mediating anthracycline-induced cardiac dysfunction.

Anthracycline-induced endotheliotoxicity has been demonstrated in various models. Wilkinson et al. demonstrated a reduction in the expression of tight junction protein, zona occludens, in doxorubicin-treated human cardiac microvascular endothelial cells, resulting in an increase in drug permeability^6^. Treatment of EA.hy926 cell line, a combination of HUVECs and A549 human lung carcinoma cell line, with anthracyclines exerting cytotoxic effects on the nuclear and cytosolic front^51^. Anthracyclines were accumulated in the nucleus in those cells even with treatment at nanomolar concentrations^51^. In addition, increase in reactive oxygen species and reorganization of F-actin cytoskeleton was observed too^51^. Another study on doxorubicin-treated HUVECs showed that protein C anticoagulant pathway is downregulated promoting a prothrombotic phenotype^52^. A doxorubicin-treated mouse model revealed greater accumulation of doxorubicin in endothelial nuclei than in other cardiac cell types ^53^. Pigs (30-35kg) treated with a cumulative dose of 2.25 mg/kg doxorubicin displayed structural damage to the arteries based on histological and myograph evaluations^54^. In children, vascular endothelial dysfunction was assessed using the brachial artery reactivity test in those who had completed anthracycline treatment at least two months prior^55^. However, their cardiac function was not reported. Altogether, these findings show that anthracyclines exert molecular, structural and functional changes to endothelial cells resulting in detrimental effects from the cellular to organ systemic levels, in various organisms. However, the condition of the endothelium at a later timepoint post-treatment has not been well characterized^11,56,57^.

This study provides a longitudinal account of the endothelial health of children with leukemia and lymphoma, by comprehensively reporting the proportion of circulating ECs and EPCs in the peripheral blood and assessing the fate of EPCs, up till 1-year post-treatment. In both leukemia and lymphoma, CECs serve as markers of endothelial damage, while EPCs are associated not only with endothelial repair but also with angiogenesis and cancer progression. Therefore, their interpretation requires careful consideration. The proportions of CECs, EPCs, and their characteristics at the pre-treatment are largely influenced by the underlying malignancy. In contrast, as participants were confirmed to be in remission from the end-of-treatment onwards. Alterations from then on were presumed to arise mainly from the residual effects of anthracycline exposure, with negligible contribution from ongoing malignancy.

### 4.1 Endothelial Health in Leukemic Patients

This study shows that in leukemia, significant endothelial injury appears to occur primarily during active malignancy which normalize during remission. However, EPC counts remain lower than in healthy controls, suggesting impaired endothelial repair capacity during remission. The finding that CEC^58^ and EPC counts are highest at initial diagnosis is consistent with another study conducted in adult AML patients^23^.

This study reveals persistent senescent EPCs during remission period. Although there is a lack of data on the fraction of senescent EPCs in leukemia patients treated with anthracyclines, a large proportion of senescent cells is expected due to the inflamed environment caused by leukemia and anthracycline treatment^59,60^. This is because inflammatory molecules activate retinoblastoma pathway triggering p16 expression inducing irreversible cell cycle arrest ^12,59,60^. The data suggests that while cancer-driven endothelial injury resolves with treatment, the persistence of senescent EPCs may hinder vascular recovery, predisposing survivors to long-term vascular dysfunction and cardiovascular complications. The impact of anthracyclines on apoptotic circulating EPCs has not been investigated in leukemia patients before. However, a previous study showed that approximately 40% of precultured primary bone marrow EPCs from adult AML patients were apoptotic before treatment ^61^. While it may seem like the proportion of apoptotic EPCs in healthy controls is high, no studies have reported this before, and findings from this study cannot be directly compared with existing data. The result may be explained by an intrinsic anti-tumour surveillance mechanism, although further studies are required to substantiate this hypothesis.

Cardiac injury based on hsTnT assay revealed significant cardiomyocyte injury and end- and 3 months post-treatment. This is corroborated by a similar study on pediatric leukemia patients treated with anthracyclines, where hsTnT levels were significantly elevated at 1-week post-treatment (11.53 pg/mL) and 6 months post-treatment (4.87 pg/mL) when compared to pre-treatment timepoint (3.50 pg/mL) ^62^. The data from this study and that of others demonstrate that delayed cardiomyocyte injury takes place in anthracycline-treated leukemia patients. However, when hsTnT concentrations were correlated with CEC counts to assess the relationship between cardiomyocyte and endothelial injury, no significant association was observed. In future studies, the use of cardiac-specific endothelial injury markers could enhance experimental robustness.

Overall, there is evidence that endothelial injury is resolved by treatment completion, while reparative capacity remains compromised in leukemia patients. This unresolved impairment could be an important driver of later vascular complications.

### 4.2 Endothelial Health in Lymphoma Patients

Just as in leukemic patients, elevated CECs may arise from either stress-induced endothelial injury or cancer-driven angiogenesis, while increased EPCs can indicate endothelial repair or cancer-associated angiogenesis^31,32^. Since participants were in remission from the end-treatment timepoint onwards, subsequent changes were not attributed to cancer progression. This study shows that lymphoma participants experience significant increase in CECs during active malignancy. This is supported by the fact that lymphoma and other cells in the microenvironment can promote angiogenesis which involves the disruption of existing blood vessels, resulting in CECs^63^. Alongside this, endothelial injury could be caused by the inflamed environment. Nevertheless, CEC levels normalized during remission, suggesting that vascular injury had subsided. Although the counts were statistically comparable to those of healthy controls, a gradual upward trend was observed from three months post-treatment onwards, suggesting the beginning of late endothelial cytotoxicity. This is plausible as a follow-up study on survivors of adolescent Hodgkin Lymphoma showed that the CEC proportion was significantly elevated even after approximately 12 years from diagnosis^64^. As for EPCs, it is no surprise that they are highest during malignancy^34,49^. While the decrease in EPCs thereafter implies remission of lymphoma and positive response to chemotherapy^49,65^, the counts decreasing below healthy levels suggests that endothelial repair is impaired.

To date, no studies have characterized EPC fate (i.e., apoptotic or senescence) in pediatric lymphoma. This study show that, like participants with leukemia, the proportion of p16⁺ EPCs in lymphoma patients is higher than in healthy controls, likely reflecting the pro-inflammatory environment induced by both the malignancy and anthracycline treatment ^60,66–68^. As observed in leukemia, the frequency of apoptotic EPCs was lower than baseline, possibly due to the opposing effects of senescence, which suppress apoptotic pathways. However, the data does show that the proportion of apoptotic EPCs increases gradually. It is possible that the levels eventually exceed those of healthy controls, as impaired EPC function has been reported in adult lymphoma patients who remained in complete remission for at least two years^69^. Lymphoma participants also exhibited the most cardiomyocyte damage at end-timepoint, which largely resolved by 3 months post-treatment. However, no significant correlation was observed between hsTnT concentrations and CEC counts.

Altogether, this study demonstrates that impairment of vascular repair mechanisms is more profound and persistent than direct vascular injury itself. This suggests that defective endothelial regeneration and persistent dysfunction in the endothelial progeny of damaged progenitor cells are key drivers of long-term vascular complications potentially leading to late-onset cardiac complications in cancer survivors^66^. This and other studies also demonstrate that apoptosis is no longer the principal mechanism underlying dox-induced endothelial injury^46^.

These findings highlight the need for therapeutic strategies aimed at restoring EPC amount and function following anthracycline treatment. The importance of EPC in vascular health is well supported by research showing that higher EPC levels are associated with a reduced risk of cardiovascular disease. EPCs can be further differentiated into “early” and “late” cells based on their functions. It is shown that early-EPCs protect themselves from apoptosis under oxidative stress^70,71^, such as anthracycline treatment through paracrine factors. While the paper primarily discusses protection against apoptosis, the interconnected nature of signaling pathways in injured cells suggests that early EPCs may also protect themselves from senescence. A related study in rats with doxorubicin-induced cardiomyopathy showed that pre-treatment with Erythropoietin reduced cardiac injury^9^. Incidentally, mice treated with Erythropoietin exhibited higher EPC counts and increased myocardial vascular density, postulating that EPCs may mediate these protective effects. Collectively, these studies strongly demonstrate the potential of EPCs as a preventive strategy against late-onset cardiac dysfunction following anthracycline treatment.

### 4.3 Limitations

While this study provides important insights, several limitations which require certain assumptions during the analysis, should be acknowledged. Firstly, we were unable to determine whether CECs originated specifically from cardiac vasculature. Therefore, it was assumed that systemic CEC fluctuations reflect cardiac endothelial alterations. To address this limitation, future work should focus on developing cardiac endothelium-specific markers ^72^. Secondly, it remains challenging to distinguish whether the EPCs are assigned for vascular repair or cancer-driven angiogenesis. Techniques such as flow cytometry, cell surface biotinylation, and mass spectrometry-based proteomics could help identify EPC subsets with unique surface markers characteristic of each process. Third, as with most cancer patients, participants in this study were administered additional medications that may influence CEC and EPC counts ^55^. Nonetheless, given the standardized treatment regimens within each type of cancer, inter-individual variability is expected to be minimal. Lastly, it was assumed that the effect of cancer is so minimal from end-timepoint onward, and thus unlikely to significantly affect the CECs and EPC counts.

## 5. Conclusion

In conclusion, this study demonstrates that impaired vascular repair during cancer remission, particularly driven by the reduced quantity and functional quality of EPCs, may underlie the increased risk of long-term cardiovascular complications in cancer survivors. Although further investigation is warranted, therapeutic and preventive strategies targeting EPC restoration and function holds promise in mitigating the cardiovascular and socioeconomic burden faced by cancer survivors.

## Data Availability

All raw data produced in the present study are available upon reasonable request to the authors.

## Abbreviations

ALL: acute lymphoblastic leukaemia
AML: acute myeloid leukaemia
BNP: brain natriuretic peptide
BSA: bovine serum albumin
CEC: circulating endothelial cells
EPC: endothelial progenitor cells
hsTnT: High-sensitivity cardiac troponin T
LVEF: left ventricular ejection fraction
NLR: Neutrophil-to-lymphocyte ratio
NT-proBNP: N-terminal pro B-type natriuretic peptide
PBS: phosphate-buffered saline
TnT: Troponin T
PBMC: peripheral blood mononuclear cell

## Perspective

### Translational Outlook

Future studies should evaluate strategies to restore endothelial progenitor cell number and function following anthracycline therapy. Targeting EPC-mediated vascular repair rather than solely preventing endothelial injury may offer a promising avenue to reduce late-onset cardiac complications in childhood cancer survivors.

### Competencies in Medical Knowledge

Further research is needed to assess whether early monitoring of EPC depletion or fate could identify individuals at elevated risk for delayed cardiac complications.

